# Conscious Sedation with Dexmedetomidine or Midazolam during Endovascular Thrombectomy in Acute Ischemic Stroke: Protocol of the Multicenter PEACE Randomized Trial

**DOI:** 10.64898/2026.09.29.26364377

**Authors:** Kang Yuan, Wusheng Zhu, Lei Yang, Xianjun Huang, Wensheng Zhang, Chunguang Chen, Jian Wang, Xiaoping Cui, Guihua Ni, Yongsheng Qiu, Shuxian Huo, Xinfeng Liu, Rui Liu, the PEACE investigators

**Author notes:** Corresponding authors: Xinfeng Liu, PhD, Department of Neurology, The First Affiliated Hospital of USTC, Division of Life Sciences and Medicine, University of Science and Technology of China, Hefei, Anhui, 230001, China., E-Mail address., Rui Liu, MD, Department of Neurology, Affiliated Jinling Hospital, Medical School of Nanjing University, No.305 East Zhongshan Road, Nanjing, 210000, Jiangsu Province, China. These authors contributed equally to this work (Kang Yuan, Wusheng Zhu).

## Abstract

**Background:** Dexmedetomidine (DEX) and midazolam (MDZ) are commonly used agents for conscious sedation in patients with acute ischemic stroke (AIS) treated with endovascular thrombectomy (EVT). While DEX offers superior sedation quality and potential neuroprotective effects, it is also associated with risk of hemodynamic instability. High-quality evidence directly comparing their impact on functional outcomes after EVT is lacking. This trial aims to determine whether conscious sedation with DEX, compared to MDZ, improves functional outcomes in patients with AIS due to anterior circulation large vessel occlusion (LVO) undergoing EVT.

**Methods:** PEACE (Dexmedetomidine versus Midazolam for Conscious Sedation during Endovascular Stroke Thrombectomy) is a prospective, randomized, open label, blinded-endpoint, multicenter trial. Up to 810 patients with AIS due to LVO in the anterior circulation within 24 hours of last known well will be randomly assigned in a 1:1 ratio to receive either DEX or MDZ for conscious sedation during EVT across 30 centers in China.

**Results:** The primary end point is the proportion of patients achieving a modified Rankin Scale (mRS) score 0-2 at 90 days, analyzed in the intention-to-treat population using a modified Poisson regression model, with results expressed as a risk ratio. Key secondary outcomes include the ordinal distribution of mRS scores at 90 days, and successful recanalization. Safety outcomes include symptomatic intracranial hemorrhage within 48 hours, and mortality at 90 days.

**Conclusion:** The PEACE trial will provide definitive, high-level evidence on the comparative efficacy and safety of DEX versus MDZ for conscious sedation during EVT in patients with AIS due to LVO in the anterior circulation, with the goal of guiding clinical practice.

**Clinical trial registration:** NCT06139692.

## Background

Stroke is the leading cause of disability and a major cause of mortality worldwide.^1^ For patients with acute ischemic stroke due to large vessel occlusion, endovascular thrombectomy (EVT) has revolutionized care and is now the standard of treatment for improving functional outcomes.^2–5^ The anesthetic management during endovascular procedure is not merely supportive but is recognized as a key modifiable factor influencing functional outcomes.^6^ While both conscious sedation and general anesthesia are utilized, conscious sedation is often preferred in clinical practice.^6,7^ Conscious sedation facilitates real-time neurological monitoring during procedures, avoids the potential hemodynamic fluctuations associated with deeper anesthesia, and is associated with faster workflow and lower costs.^8^

The choice of sedative agent during conscious sedation, however, presents a common and consequential clinical dilemma. Dexmedetomidine (DEX) and midazolam (MDZ) are the most commonly used agents, yet they have distinct pharmacological and risk-benefit profiles. MDZ, a conventional benzodiazepine, provides reliable sedation but may be associated with respiratory depression and delayed neurological assessment.^9^ In contrast, DEX, a highly selective α2-adrenergic agonist, provides a unique “cooperative sedation” that facilitates patient interaction.^10^ Importantly, beyond its sedative properties, preclinical and some clinical data suggest that DEX may confer neuroprotective benefits,^11^ such as stabilizing the blood–brain barrier and attenuating the inflammatory and oxidative stress responses to ischemia.^12–14^ Observational studies have further linked its use to a potential improvement in functional recovery after EVT.^15–17^ Despite these promising insights, a direct comparison of the two agents within a randomized trial is lacking. This issue is important because the potential advantages of DEX are balanced against significant concerns; its use can provoke hemodynamic instability, notably bradycardia and hypotension, which could in turn exacerbate cerebral injury within the ischemic penumbra.^18,19^ Consequently, current clinical guidelines have not yet provided specific recommendations on sedative choice, leaving practitioners without high-level evidence to inform this routine yet high-stakes decision.

Therefore, the PEACE (Dexmedetomidine versus Midazolam for Conscious Sedation during Endovascular Stroke Thrombectomy) randomized trial is designed and conducted to determine the efficacy and safety of DEX versus MDZ for conscious sedation during EVT in acute ischemic stroke patients secondary to large vessel occlusion in the anterior circulation.

## Methods

### Data availability

The data that support the findings of this study are available on reasonable request from the corresponding authors.

### Design

The PEACE trial is a multicenter, randomized, open label trial with blinded assessment of endpoints, aiming to investigate the efficacy and safety of DEX versus MDZ for conscious sedation during EVT in patients with acute ischemic stroke due to large vessel occlusion within 24 hours of last known well across 30 centers in China. The participant flow is illustrated in **Figure 1**. The trial is designed in compliance with the Declaration of Helsinki and has been registered at https://clinicaltrials.gov (identifier: NCT06139692). The protocol has been approved by the Clinical Trials Ethics Committee of Jinling Hospital and all participating hospitals prior to enrollment.

**Figure 1.**
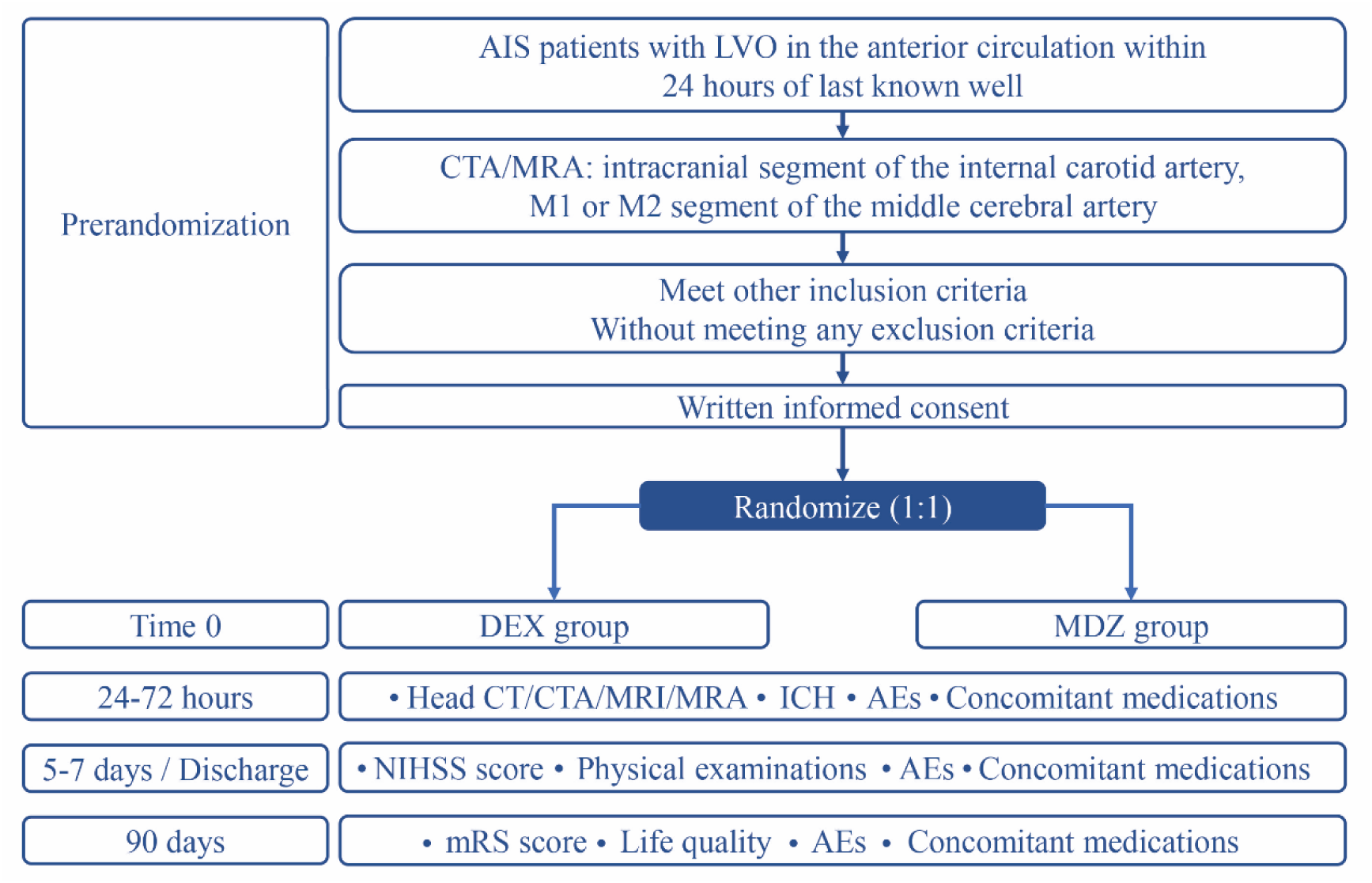
Flow chart. Abbreviations: AEs, adverse events; AIS, acute ischemic stroke; CT, computed tomography; CTA, computed tomographic angiography; DEX, dexmedetomidine; ICH, intracerebral hemorrhage; LVO, large vessel occlusion; MDZ, midazolam. MRA, magnetic resonance angiography; MRI, magnetic resonance imaging; mRS, modified Rankin Scale; NIHSS, National Institutes of Health Stroke Scale.

**Table 1.**
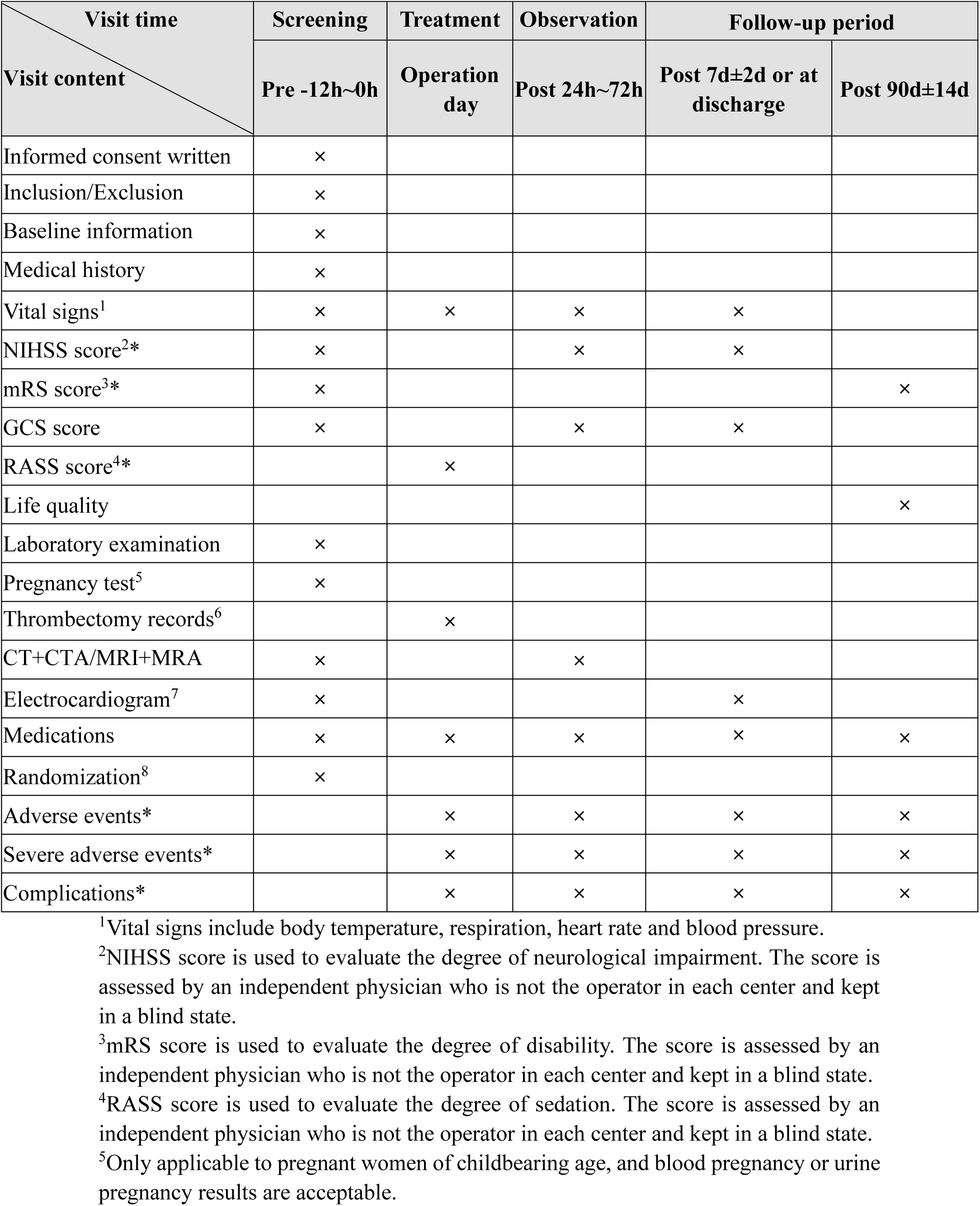
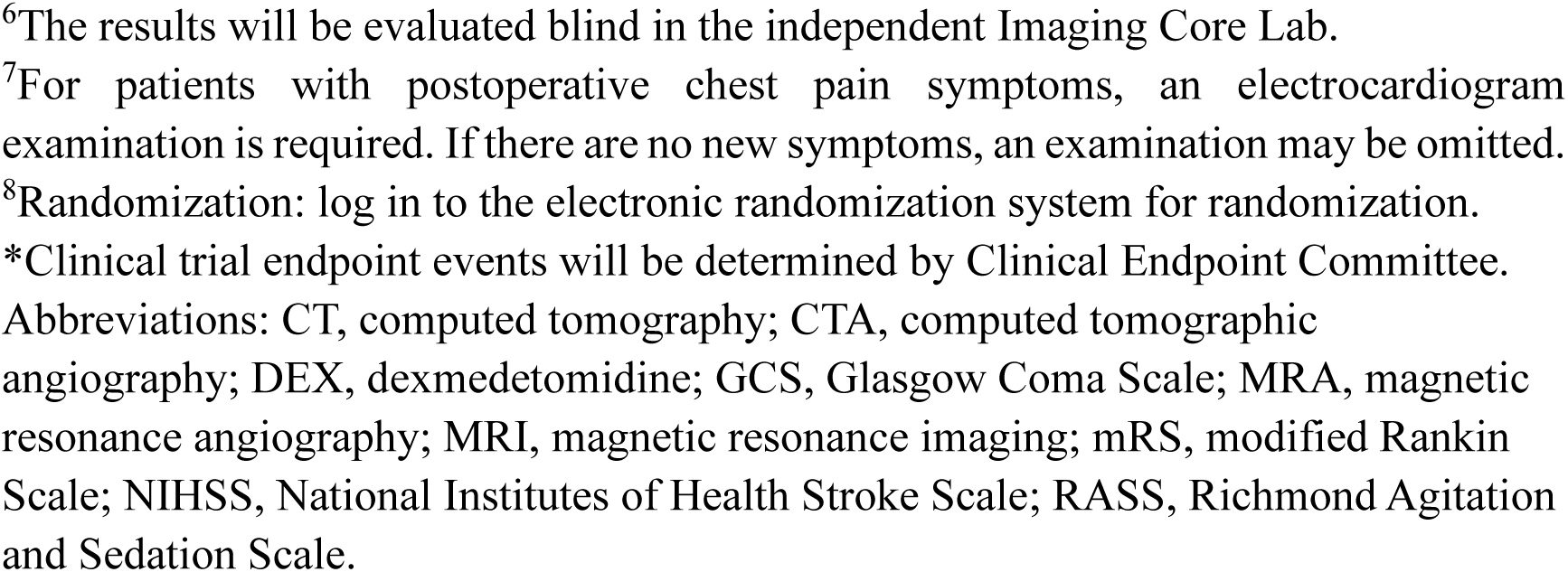
Trial schedule.

| Visit time<br>Visit content | Screening | Treatment | Observation | Follow-up period |  |
| --- | --- | --- | --- | --- | --- |
|  | Pre -12h~0h | Operation day | Post 24h~72h | Post 7d±2d or at discharge | Post 90d±14d |
| Informed consent written | × |  |  |  |  |
| Inclusion/Exclusion | × |  |  |  |  |
| Baseline information | × |  |  |  |  |
| Medical history | × |  |  |  |  |
| Vital signs<sup>1</sup> | × | × | × | × |  |
| NIHSS score<sup>2*</sup> | × |  | × | × |  |
| mRS score<sup>3*</sup> | × |  |  |  | × |
| GCS score | × |  | × | × |  |
| RASS score<sup>4*</sup> |  | × |  |  |  |
| Life quality |  |  |  |  | × |
| Laboratory examination | × |  |  |  |  |
| Pregnancy test<sup>5</sup> | × |  |  |  |  |
| Thrombectomy records<sup>6</sup> |  | × |  |  |  |
| CT+CTA/MRI+MRA | × |  | × |  |  |
| Electrocardiogram<sup>7</sup> | × |  |  | × |  |
| Medications | × | × | × | × | × |
| Randomization<sup>8</sup> | × |  |  |  |  |
| Adverse events* |  | × | × | × | × |
| Severe adverse events* |  | × | × | × | × |
| Complications* |  | × | × | × | × |

<sup>1</sup>Vital signs include body temperature, respiration, heart rate and blood pressure.
<sup>2</sup>NIHSS score is used to evaluate the degree of neurological impairment. The score is assessed by an independent physician who is not the operator in each center and kept in a blind state.
<sup>3</sup>mRS score is used to evaluate the degree of disability. The score is assessed by an independent physician who is not the operator in each center and kept in a blind state.
<sup>4</sup>RASS score is used to evaluate the degree of sedation. The score is assessed by an independent physician who is not the operator in each center and kept in a blind state.
<sup>5</sup>Only applicable to pregnant women of childbearing age, and blood pregnancy or urine pregnancy results are acceptable.
<sup>6</sup>The results will be evaluated blind in the independent Imaging Core Lab.
<sup>7</sup>For patients with postoperative chest pain symptoms, an electrocardiogram examination is required. If there are no new symptoms, an examination may be omitted.
<sup>8</sup>Randomization: log in to the electronic randomization system for randomization.
\*Clinical trial endpoint events will be determined by Clinical Endpoint Committee.
Abbreviations: CT, computed tomography; CTA, computed tomographic angiography; DEX, dexmedetomidine; GCS, Glasgow Coma Scale; MRA, magnetic resonance angiography; MRI, magnetic resonance imaging; mRS, modified Rankin Scale; NIHSS, National Institutes of Health Stroke Scale; RASS, Richmond Agitation and Sedation Scale.

### Patient population

#### Inclusion criteria

1. Age ≥ 18 years;
2. Presenting with symptoms of acute ischemic stroke within 24 hours of last known well;
3. Computed tomography angiography or magnetic resonance angiography confirmed occlusion of the intracranial segment of the internal carotid artery, M1 or M2 segment of the middle cerebral artery;
4. Pre-stroke modified Rankin scale (mRS) score ≤ 2;
5. National Institutes of Health Stroke Scale (NIHSS) score ≥ 6 before randomization;
6. Alberta Stroke Program Early CT Score (ASPECTS) ≥ 3 prior to randomization;
7. The patient or their legal representative provides written informed consent.

#### Exclusion criteria

1. Pregnant or lactating women;
2. Known allergy to contrast agents or nitinol devices;
3. Known allergy to midazolam or other benzodiazepines;
4. Known allergy to dexmedetomidine or its components;
5. Long-term use of sedative medications, α2-adrenergic receptor agonists (such as clonidine), or potent CYP450 inhibitors or inducers before randomization;
6. Uncontrolled hypertension or hypotension (defined as systolic blood pressure > 185 mmHg or < 90 mmHg, diastolic blood pressure > 110 mmHg or < 60 mmHg) measured prior to randomization;
7. Second-degree or third-degree heart blockage or bradyarrhythmia with a baseline heart rate lower than 50 beats / min;
8. Any major surgery or serious trauma within 14 days;
9. Known genetic or acquired bleeding diathesis (platelet count < 100*10^9/L, activated partial thromboplastin time > 50 s or international normalized ratio > 1.7);
10. Blood glucose < 2.8 or > 22.2 mmol/L;
11. Severe renal insufficiency (defined as glomerular filtration rate < 30 ml/min or serum creatinine > 220 mmol/L [2.5mg/dl]);
12. Receiving hemodialysis or peritoneal dialysis;
13. Life expectancy ≤ 1 year;
14. Severe agitation or seizures;
15. Clinical manifestations of central nervous system vasculitis;
16. Premorbid neurological disease or mental disorders confounding evaluation;
17. Unwilling to be followed up within 90 days;
18. Participation in other interventional randomized clinical trials.

#### Imaging exclusion criteria

1. Evidence of intracranial hemorrhage on computed tomography (CT) or magnetic resonance imaging (MRI);
2. Cerebellar infarction with obvious space-occupying effects and fourth ventricle compression on CT or MRI;
3. Any untreated or incompletely treated intracranial aneurysm or any intracranial vascular malformation;
4. Bilateral occlusion or multiple intracranial vessels occlusions;
5. Intracranial tumors (with mass effect).

### Randomization and masking

The PEACE trial employs a stratified block randomization method for randomization. Patients will be stratified based on the following three factors: 1) age (≤70 vs. >70 years), 2) sex (female vs. male), and 3) time from last known well time to hospital arrival (≤ 120 vs. > 120 minutes). A central, web-based randomization system (Interactive Web Response System [IWRS]) will be employed to manage the allocation process. A computer-generated randomization sequence, prepared using SAS 9.4 software in a 1:1 ratio (DEX vs. MDZ), will be loaded into the system. Upon enrollment of an eligible patient, the neuro-interventionalists and anesthesiologists who are administering the sedating agent will access the IWRS to obtain the immediate treatment assignment, ensuring concealment of the upcoming allocation.

Given the inherent differences in drug administration between the two intervention arms, blinding of the neuro-interventionalists and anesthesiologists who are administering the sedating agent is not feasible. Therefore, the trial is open-label at the point of care. However, to minimize bias in outcome assessment, a single-blind design will be adopted, whereby investigators and all study personnel involved in post-procedural management, data collection, and clinical follow-up will remain blinded to treatment allocation. The treatment allocation will be strictly concealed from the patient, outcomes assessors (such as those performing the 90-day mRS assessment), and the biostatisticians performing the primary analysis. Access to the unblinded allocation will be restricted exclusively to the designated procedural personnel, who will be instructed not to disclose this information.

### Treatments

The assigned sedative agent will be initiated at the beginning of the EVT procedure and titrated to achieve a level of conscious sedation considered appropriate by the neuro-interventionalists to ensure safe and effective procedure execution. The specific protocols for each group are detailed below. Investigators will be advised to maintain systolic blood pressure <180 mmHg before and during EVT and within a target range of 140–180 mmHg after the procedure. Antihypertensive or vasopressor agents may be administered as clinically indicated according to institutional protocols.

Hemodynamic parameters, including heart rate and blood pressure, will be continuously monitored during the procedure and at each follow-up visit. If clinically significant pain occurs after EVT, analgesics (e.g., fentanyl) can be administered as clinically indicated.

#### DEX group

For patients allocated to the DEX group, sedation will be induced with an intravenous loading dose of 1.0 μg/kg administered over 10 minutes, prepared from a standard 8 μg/mL solution. This will be immediately followed by a continuous maintenance infusion, delivered via pump at a rate ranging from 0.2 to 0.7 μg/kg/h, with adjustments made according to the desired clinical effect. If adequate sedation cannot be achieved with DEX alone, physicians or anesthesiologists are permitted to administer rescue sedation agents (such as propofol) or consider transitioning the patient to general anesthesia.

#### MDZ group

For patients allocated to the MDZ group, sedation will be initiated with an intravenous bolus of 0.05 mg/kg, drawn from a standard 1 mg/mL solution. A continuous maintenance infusion will then be started at a rate between 0.04 and 0.2 mg/kg/h, titrated to effect using an infusion pump. The infusion rate can be adjusted based on the sedation effect to achieve the appropriate level of sedation during EVT. If MDZ does not achieve satisfactory sedation during procedure, physicians or anesthesiologists may choose rescue sedation agents (such as propofol) or consider transitioning the patient to general anesthesia.

### Efficacy end points

#### Primary end point

The primary end point was the proportion of mRS score 0-2 at 90 days. The mRS score has a minimum value of 0 and maximum value of 6. Higher value indicated worse functional outcome.

#### Secondary efficacy end points

1. Ordinal distribution of mRS score at 90 days;
2. Recanalization rate on final angiography of EVT defined as expanded Thrombolysis in Cerebral Infarction score of 2b-3;
3. Score on the ASPECTS at 24-72 hours assessed with CT / MRI;
4. Deep sedation defined as Richmond Agitation and Sedation Scale (RASS) score ≤ -3;
5. Score on the Glasgow Coma Scale (GCS) at 5-7 days;
6. Score on the NIHSS at 5-7 days;
7. Score on the Barthel Index at 90 days;
8. Score on the EQ-5D-5L at 90 days.

#### Tertiary efficacy end points

1. Proportion of mRS score 0-1 at 90 days;
2. Proportion of mRS score 0-3 at 90 days;
3. Under sedation defined as RASS score ≤ 0;
4. Score on the GCS at 24 hours;
5. Score on the NIHSS at 24 hours;

#### Safety end points

1. Proportion of symptomatic intracranial hemorrhage at 48 hours; Symptomatic intracranial hemorrhage is assessed using the Heidelberg Bleeding Classification;
2. Proportion of procedure-related complications;
3. Proportion of mortality at 90 days;
4. Proportion of adverse events;
5. Proportion of severe adverse events.

### Data and safety monitoring board (DSMB)

An independent DSMB will be established, comprising three experts in the fields of neurology, neuro-intervention, and biostatistics, respectively. All members are independent of the study sponsors and are not involved in the conduct of the trial. The DSMB will convene annually to monitor the progress of the trial and will also review the incidence of serious adverse events to ensure patient safety throughout the study. The DSMB will have unblinded access to all safety and efficacy data. Based on its reviews, the board is empowered to make formal, confidential recommendations to the trial Steering Committee regarding the continuation, modification, or termination of the study.

### Sample size estimate

Previous studies reported that the proportion of favorable functional outcome (mRS score 0-2) at 90 days under conventional sedation is approximately 50% in acute ischemic stroke patients with large vessel occlusion in the anterior circulation.^15^ We hypothesize that sedation with DEX may improve this outcome, corresponding to an absolute increase of 10% (from 40% to 50%, representing a 25% relative increase). With a two-sided significance level of α = 0.05 and a statistical power of 80%, the required sample size to detect this difference is 770 patients (385 per group). To account for an estimated 5% loss to follow-up or dropout, the total enrollment target is increased to 810 patients (405 per group). This estimation was performed using PASS software (NCSS, LLC, Kaysville, USA) version 15.0.

### Statistical analyses

The primary efficacy analysis will be performed in the intention-to-treat population, which includes all randomized patients according to their assigned treatment group. Safety analyses will be performed on the safety dataset, comprising all patients who received at least part of the study intervention.

For the primary endpoint, the proportion of patients with the mRS score of 0–2 at 90 days, the comparison between the DEX and MDZ groups will be conducted using a modified Poisson regression model. Results will be reported as risk ratio with corresponding 95% confidence interval. The PEACE trial incorporates two planned interim analyses for efficacy. These will be conducted by the independent DSMB after approximately 1 / 3 (n = 270) and 2 / 3 (n = 540) of the total sample size have completed the 90-day follow-up. The O’Brien-Fleming method will be applied for alpha adjustments (first analysis α=0.00021, second analysis α=0.01202, final analysis α=0.04626).

Secondary outcomes (e.g., ordinal shift in mRS scores, successful recanalization, quality of life scores) and tertiary outcomes will be analyzed using ordinal logistic regression, linear regression, or modified Poisson regression, as appropriate. Missing data for the primary outcome will be imputed using the multiple imputation. Missing values for baseline characteristics as well as secondary and tertiary outcomes will be handled using multiple imputation methods.

In addition, subgroup analyses will be conducted to explore the consistency of the treatment effect across clinically relevant subgroups. Prespecified subgroups include: intravenous thrombolysis (yes vs. no), wake-up stroke (yes vs. no), baseline ASPECTS (3-6 vs. 6-10), baseline NIHSS score (> 16 vs. ≤ 16), occlusion site (internal carotid artery vs. middle cerebral artery), stroke etiology (large artery atherosclerosis vs. cardioembolism vs. other / undetermined causes), and stroke sidedness (left vs. right).

All statistical analyses will be conducted using R software (version 4.2.1, R Core Team, Vienna, Austria). A two-sided *P* value < 0.05 will be considered statistically significant for the final analysis of the primary endpoint, respecting the adjusted alpha boundaries for interim looks.

## Discussion

The PEACE trial is a multicenter, randomized trial designed to investigate an important and unresolved clinical issue in EVT for acute ischemic stroke: which sedative agent, DEX or MDZ, is superior for conscious sedation during EVT procedure in patients with acute ischemic stroke due to large vessel occlusion in the anterior circulation within 24 hours of last known well.

While DEX may provide neuroprotective benefits, and has been associated with more stable intraprocedural hemodynamics and potentially improved functional outcomes in several observational studies,^15^ other analyses have linked its use to a higher incidence of hypotension, bradycardia, and worse functional recovery.^20^ This controversy is reflected in the absence of specific recommendations in clinical guidelines, leaving the choice of sedative agent to institutional preference or individual clinician judgment. The PEACE trial may elucidate the efficacy of DEX and MDZ for conscious sedation during EVT and their effects on functional outcomes, providing evidence to achieve optimal sedation efficiently, expedite vessel recanalization, and enhance recovery in acute ischemic stroke patients.

The design of the PEACE trial is methodologically tailored to provide a definitive answer to this clinical dilemma. First, to ensure broad generalizability, the inclusion criteria are aligned with contemporary EVT guidelines, enrolling a representative study population of patients with anterior circulation large vessel occlusion. Second, key safety exclusions, such as pre-existing bradycardia, severe hypotension, and known drug allergies, are implemented to mitigate the specific risks associated with each sedative, thereby isolating their comparative efficacy under optimized conditions. Third, the protocol mandates the use of conscious sedation, reflecting its status as the preferred technique in most high-volume stroke centers due to advantages in workflow and neurological monitoring. Additionally, although EVT may be performed in selected patients with coagulopathy in clinical practice, patients with significant bleeding diathesis were excluded from the present study because of the potential risk of hemorrhagic complications, to ensure patient safety and minimize potential confounding related to severe coagulopathy.

A critical design consideration is the open-label nature of the intervention, necessitated by the distinct administration protocols for DEX and MDZ. To minimize assessment bias, rigorous blinding procedures are enforced for outcome assessors, follow-up personnel, and statisticians. Furthermore, the trial incorporates a pre-specified statistical analysis plan, two interim analyses by an independent DSMB, and adjustment for stratification factors, all of which strengthen the validity of the final conclusions.

To our knowledge, only one ongoing randomized controlled trial—the PPDET trial (NCT04916197)—is investigating the impact of prolonged use of DEX after EVT on long-term prognosis of acute ischemic stroke patients.^21^ Different from the PPDET trial, the present trial focuses on comparing DEX and MDZ for conscious sedation during EVT. If DEX shows superior efficacy in improving 90-day functional outcomes without an elevated risk of intracranial hemorrhage or mortality, it could establish a new standard of care. Conversely, if no significant difference or a safety concern emerges, it would validate the continued use of midazolam as a safe and effective alternative.

The PEACE trial enrolled its first patient on November 20, 2023, and is ongoing. As of manuscript submission, 752 patients have been recruited. Enrollment will be ended in April 2026, with 90-day follow-up anticipated to be completed by July 2026.

## Conclusion

The PEACE multicenter randomized trial comparing DEX and MDZ for conscious sedation during EVT in patients with acute ischemic stroke due to large vessel occlusion in the anterior circulation. Designed with pragmatic inclusion criteria and rigorous methodology, it aims to provide definitive evidence on which sedative leads to better 90-day functional outcomes, guiding clinical practice and future guidelines.

## Data Availability

All data produced in the present study are available upon reasonable request to the authors

## Acknowledgments and declaration of conflicting interest

None.

## Nonstandard Abbreviations and Acronyms

ASPECTS: Alberta Stroke Program Early CT Score
CT: Computed tomography
DEX: Dexmedetomidine
DSMB: Data and safety monitoring board
EVT: Endovascular thrombectomy
GCS: Glasgow Coma Scale
MDZ: Midazolam
MRI: Magnetic resonance imaging
mRS: modified Rankin Scale
NIHSS: National Institutes of Health Stroke Scale
PEACE: Dexmedetomidine versus Midazolam for Conscious Sedation during Endovascular Stroke Thrombectomy
RASS: Richmond Agitation and Sedation Scale
IWRS: Interactive Web Response System

